# coronabambini.ch: Development and usage of an online decision support tool for pediatric COVID-testing in Switzerland: a cross-sectional analysis

**DOI:** 10.1101/2022.12.19.22283560

**Authors:** C Starvaggi, N Travaglini, C Aebi, F Romano, I Steiner, T C Sauter, K Keitel

## Abstract

**Objectives:** To describe the development and usage of coronabambini.ch as an example of a pediatric electronic public health application and to explore its potential and limitations in providing information on disease epidemiology and public health policy implementation.

**Design:** We developed and maintained a non-commercial online decision support tool, coronabambini.ch, to translate the Swiss FOPH pediatric (age 0-18 years) COVID-19 guidelines around testing and school/daycare attendance for caregivers, teachers, and healthcare personnel. We analyzed the online decision tool as well as a voluntary follow-up survey from October 2020 to September 2021 to explore its potential as a surveillance tool for public health policy and epidemiology.

**Participants:** 68’269 users accessed and 52’726 filled out the complete online decision tool. 3% (1’399/52’726) filled out a voluntary follow-up. 92% (18’797/20’330) of users were parents.

**Results:** Certain dynamics of the pandemic and changes in testing strategies were reflected in the data captured by coronabambini.ch: e.g. in terms of disease epidemiology, gastrointestinal symptoms were reported more frequently in younger age-groups (13% (3’308/26’180) in children 0-5 years versus 9% (3’934/42’089) in children ≥6 years, 𝒳^2^=184, *P*=<.001). As a reflection of public health policy, the proportion of users consulting the tool for a positive contact without symptoms in children 6-12 years increased from 4% (1’415/32’215) to 6% (636/9’872) after the FOPH loosened testing criteria in this age-group, 𝒳^2^=69, *P=*<.001. Adherence to the recommendation was generally high (84% (1’131/1’352)) but differed by the type of recommendation: 89% (344/385) for “stay at home and observe”, 75% (232/310) for “school attendance”.

**Conclusions:** Usage of coronabambini.ch was generally high in areas where it was developed and promoted. Certain patterns in epidemiology and adherence to public health policy could be depicted but selection bias was difficult to measure showing the potential and challenges of digital decision support as public health tools.

**Strengths and limitations of this study:**

- We developed and maintained a non-commercial online decision support tool, coronabambini.ch, based on the Swiss Federal Office of Public Health (FOPH) pediatric COVID-19 guidelines around testing and school/daycare attendance.
- We performed a descriptive analysis of the usage data from the tool between October 2020 to September 2021 to explore its potential as a surveillance tool for public health policy and epidemiology.
- A follow-up survey measured comprehensibility and adherence of the online decision tool.
- The types of selection biases of the data were difficult to assess within this voluntary online survey system.
- Response rate of the follow-up survey was low and participants with high education and high health literacy were overrepresented.

## Introduction

The Swiss Federal Office of Public Health (FOPH) published the first national pediatric COVID-19 testing guidelines in June 2020 [1]. The testing algorithm was developed in conjunction with the national association of pediatrics and intended to guide parents and healthcare providers to decide which children should be tested for COVID-19 and regarding school/daycare attendance. Special focus was laid on i) minimizing testing burden on children <12 years of age, ii) allowing school and daycare attendance as much as possible, iii) strengthening the role of pediatric primary care providers as gatekeepers, and iv) avoiding changes in care-seeking behavior when children are acutely ill. The resulting testing guideline was elaborate but also complex to interpret for laypersons as well as for healthcare providers.

To support the implementation of these pediatric COVID-19 testing guidelines we developed an online decision support tool in collaboration with the FOPH, coronabambini.ch [2]. Such a decision tool may provide a current, credible and practical source of information for caregivers and health care personnel in decisions around testing and attendance of school and daycare. Decision support tools to support public health policies, rather than medical decision support tools, are relatively new. However, during the current COVID-19 pandemic several online triage decision tools for adults have been developed to relieve stretched healthcare systems [3-13]. The requirements for such online decision supports tools in terms of timeliness and flexibility in adapting to changing guidelines are high and represent a major challenge in operating such an instrument [3]. Tools for children are even more challenging as they have to consider various user groups and more complex guidelines. The potential of such online tools may go beyond decision support as they could serve as surveillance tools for disease epidemiology and policy implementation [7-9,13,14]. However, important and inherent methodological limitations include selection bias, such as an overrepresentation of users with high education status.

The objective of this manuscript is to describe the development and usage of coronabambini.ch as an example of a pediatric electronic public health application and to explore its potential and challenges in providing information on disease epidemiology and public health policy implementation.

## Methods

### Study Design

This was a descriptive analysis of data generated by the online decision tool (coronabambini.ch) as well as a voluntary follow-up survey from October 2020 to September 2021 to explore its potential as a surveillance tool for public health policy and epidemiology.

### Development of the online decision support tool

We developed and maintained an online decision support tool as a new non-commercial initiative based on the official pediatric testing guidelines published by the FOPH.

The pediatric COVID-19 testing guidelines were translated into a binary decision tree in collaboration with members of the Swiss pediatric COVID-19 expert group and the FOPH (Figure S1). The initial FOPH version of the guideline recommended different testing criteria for children <12 years. For children 12 years and older the same testing criteria as for adults were applied [1]. We split the age group of <12 years into three age groups: <3 months, 3 months to 5 years and 6 to 11 years. This allowed to accommodate different advice for higher risk groups (e.g. to consult a pediatrician the same day in case of fever for children <3 months) and to consider age-appropriate symptoms (e.g. loss of smell only for older children). In March 2021 the FOPH pediatric testing strategy changed in that testing according to adult criteria was recommended for children 6 years and above [1]. The online decision tool was adapted accordingly shortly thereafter.

The algorithm based on the decision tree was then programmed into an online survey tool (onlineumfragen.com) [15]. The tool consists of two parts: mandatory algorithmic entry fields, which are required to provide users with the testing recommendation, followed by a voluntary section covering demographic information. At the end, users were asked to voluntarily provide their e-mail address for a follow-up survey. Table S1 displays the content of the online decision support tool and the voluntary questions at the end of the tool. Sample screenshots are shown in Figure S2.

### Participants, setting, data sources and measurements

After internal validation, a first version of the tool was published on October 15, 2020 (Figure S1) and was openly accessible for users across Switzerland. As of the 03.12.2020 it was fully integrated in the online decision support tool for adults developed by the FOPH. The tool was also advertised through the Swiss Pediatric Society. Target users (and hence participants) included caregivers and teachers. After the Swiss Federal Council lifted most of the restrictions that were put in place during the pandemic on February 16, 2022, the FOPH’s online decision support tool for adults as well as coronabambini.ch were discontinued. For this analysis, we considered entries over 12 months from October 2020 to September 2021, which corresponded to the second and third COVID-waves in Switzerland. It also considers entries from the beginning of the fourth COVID-wave (delta-variant) starting in the late summer of 2021.

Users of the online decision support tool who voluntarily provided an email address received an automatic e-mail notification with a follow-up survey 14 days after the initial survey (Figure 1). The follow-up contained questions around the reasons for usage, demographic and socio-economic information, as well as adherence to the testing recommendation (Table S2).

**Figure 1.**
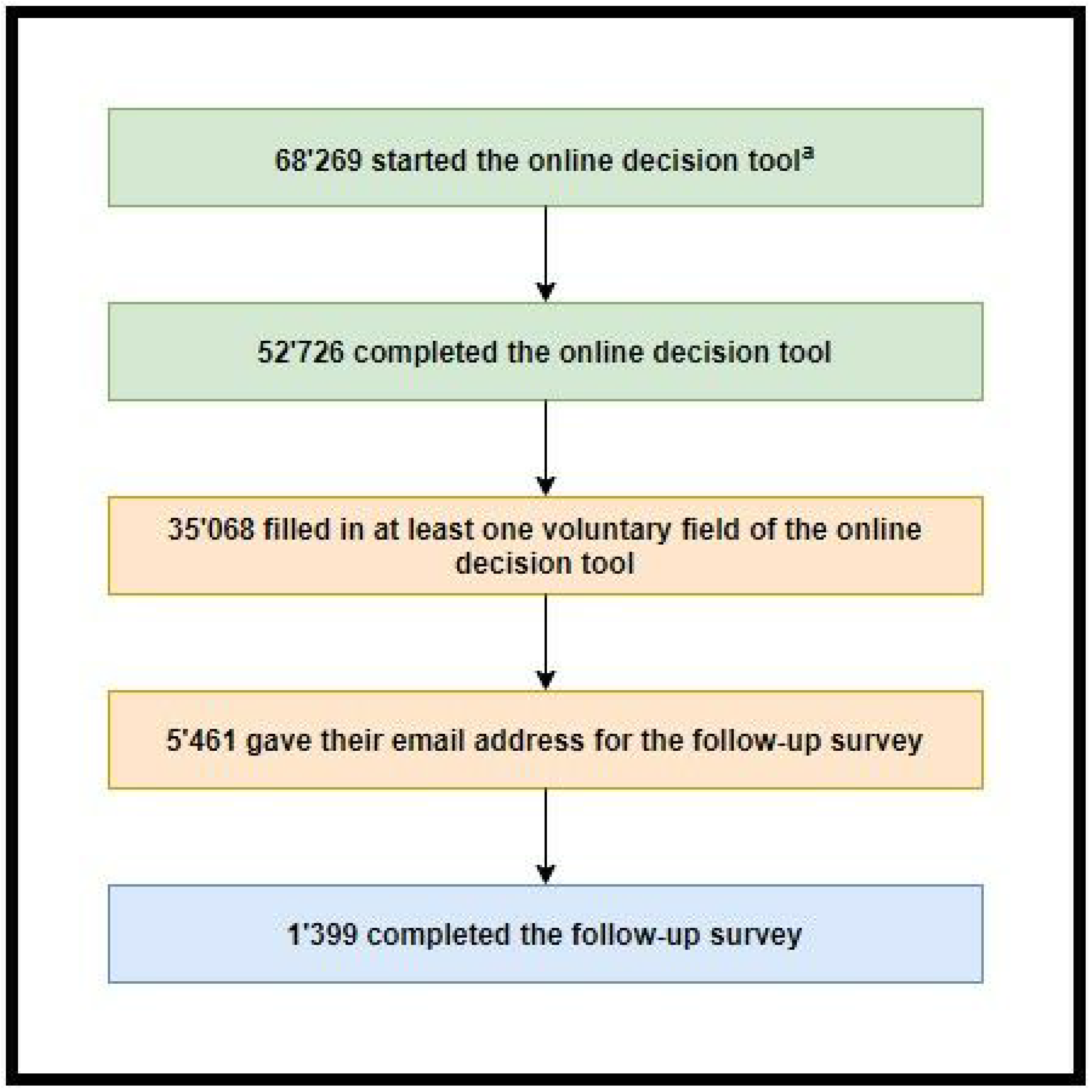
Flow-chart of the usage of coronabambini.ch: The online decision tool consists of mandatory fields (green) that are necessary to provide the testing recommendations. The mandatory section is followed by a voluntary section containing fields on demographics and users are asked to provide their e-mail address for the follow-up survey (blue). The follow-up survey is sent automatically to the email addresses provided after 14 days. ^a^ A form was opened when at least one question was answered

### Statistical Methods

Data analysis was performed using Stata^®^MP 16 (64-bit) (StataCorp, The College Station, Texas, USA). For categorical variables frequency and percentages were calculated for the descriptive analysis. Differences in the groups of interest were assessed by using the Pearson Chi-square test. *P* <.05 was considered significant. Regarding the voluntary fields, only those with attributable answers were considered.

### Patient and public involvement

Patients and the public were not involved in the design, conduct, reporting, or dissemination plans of this research.

## Results

### Usage

From October 2020 to September 2021, 68’269 users answered at least one question including 52’726 (77% (52’726/68’269)) answering all mandatory questions of the online decision tool (Figure 1). 5’461 users provided their email address for the voluntary follow-up survey. 25.6% (1’399/5’461) filled in the follow-up survey. Figure 2 shows the numbers of visits in relation to the COVID-19 case numbers in Switzerland during the data collection period. The most selected age-category were children aged 6 – 12 years (55% (27’803/50’896)) before the change in testing strategy, and children aged ≥6 years (57% (9’874/17’373) after April 2021.

**Figure 2.**
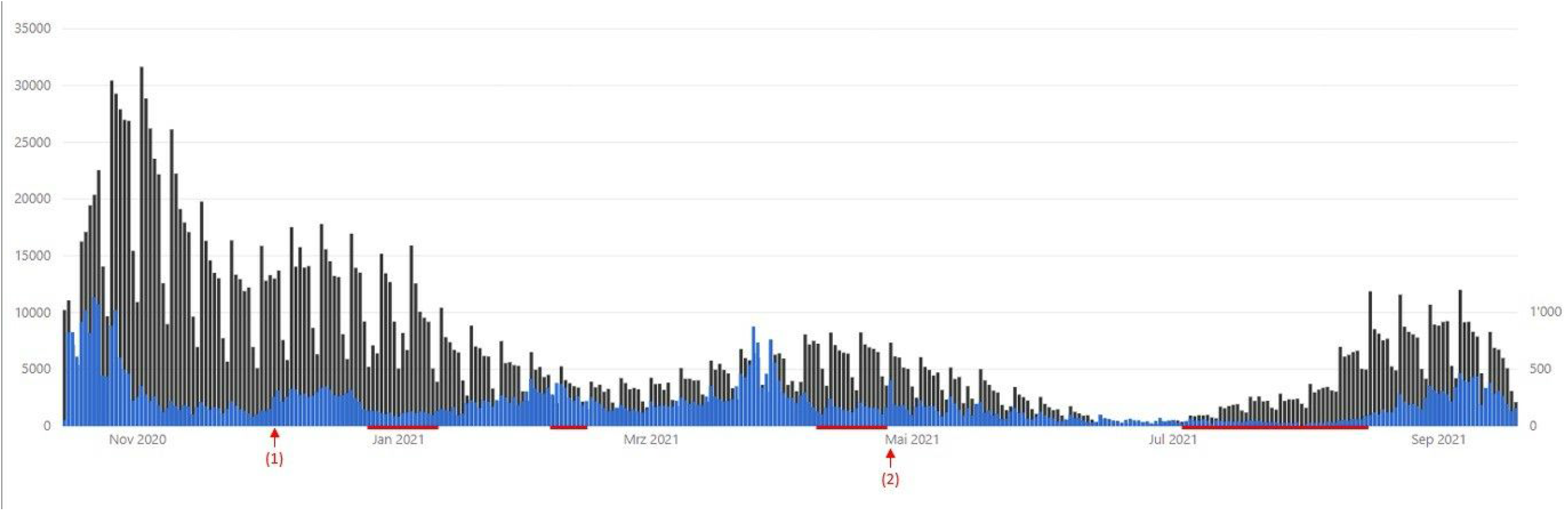
Use of the online decision tool (right scale, blue) compared to the reported cases by the FOPH (left scale, black). (1) marks the day that coronabambini.ch was integrated to the COVID-checker of the FOPH. (2) marks the day the algorithm changed. The red bars mark the school-holidays of the city of Bern.

### Mandatory fields for testing recommendation

Age category, symptoms, contact to a confirmed or suspected case, and contacted by the cantonal contact tracing were mandatory fields and are shown in Table 1. Upper respiratory symptoms were the most common symptoms in all age groups (Table 1). Gastrointestinal symptoms (diarrhea and or vomiting) were more frequently reported in the younger age groups, when compared to older children. 16% (10’661/68’269) of users reported no symptoms. We did not note relevant differences in symptom reporting between parents (mother or father) and other users (Table S3).

**Table 1.**
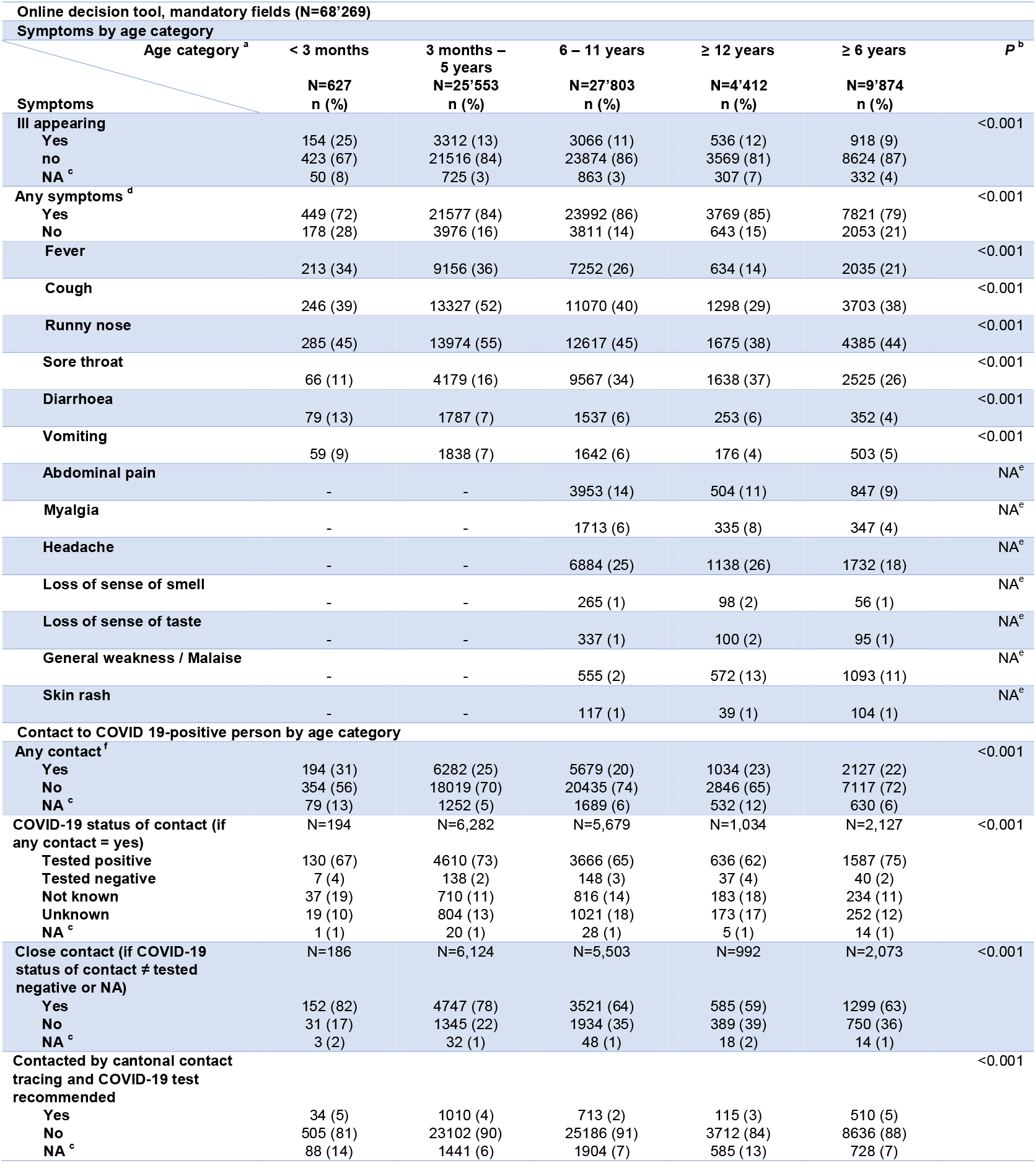
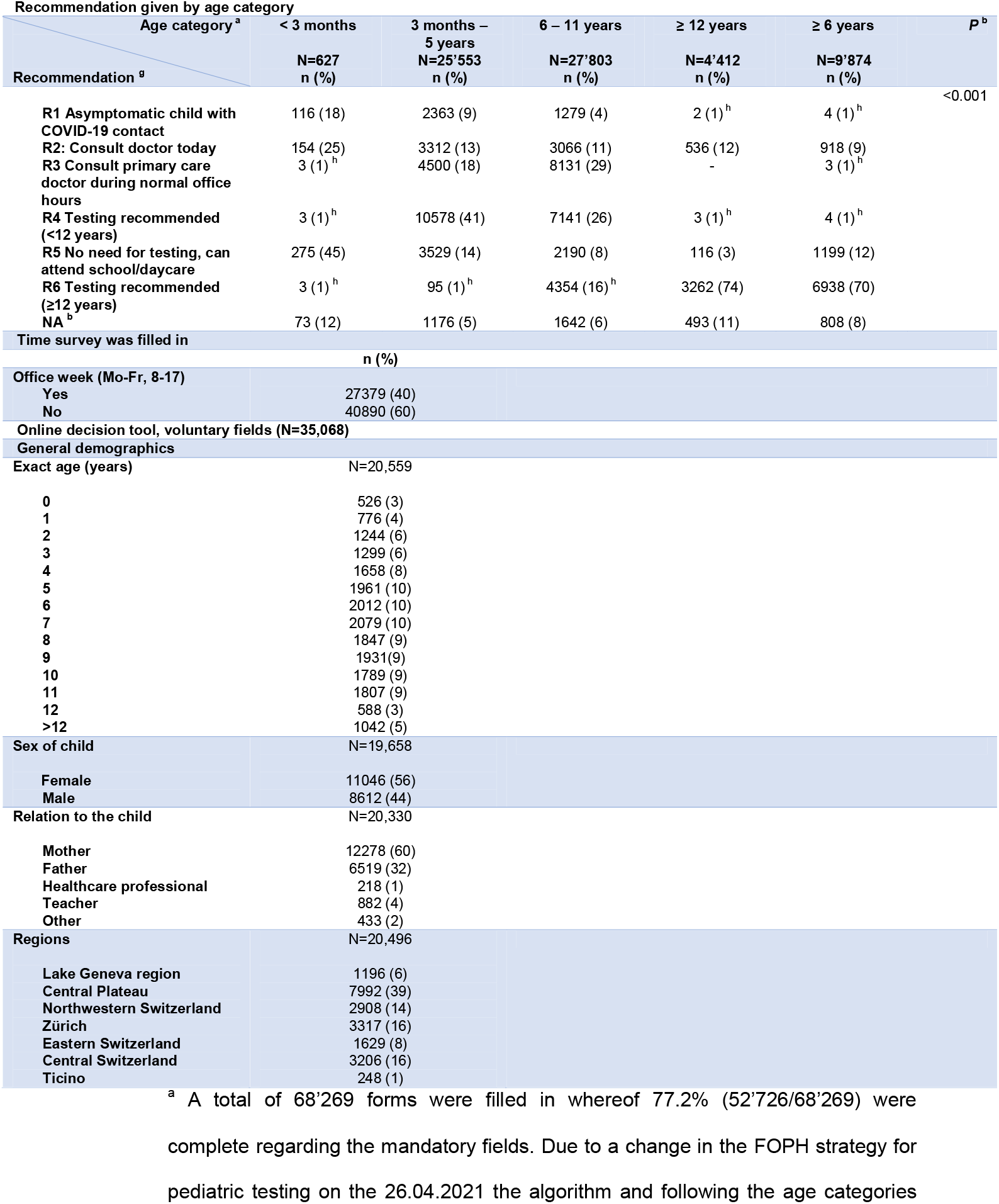
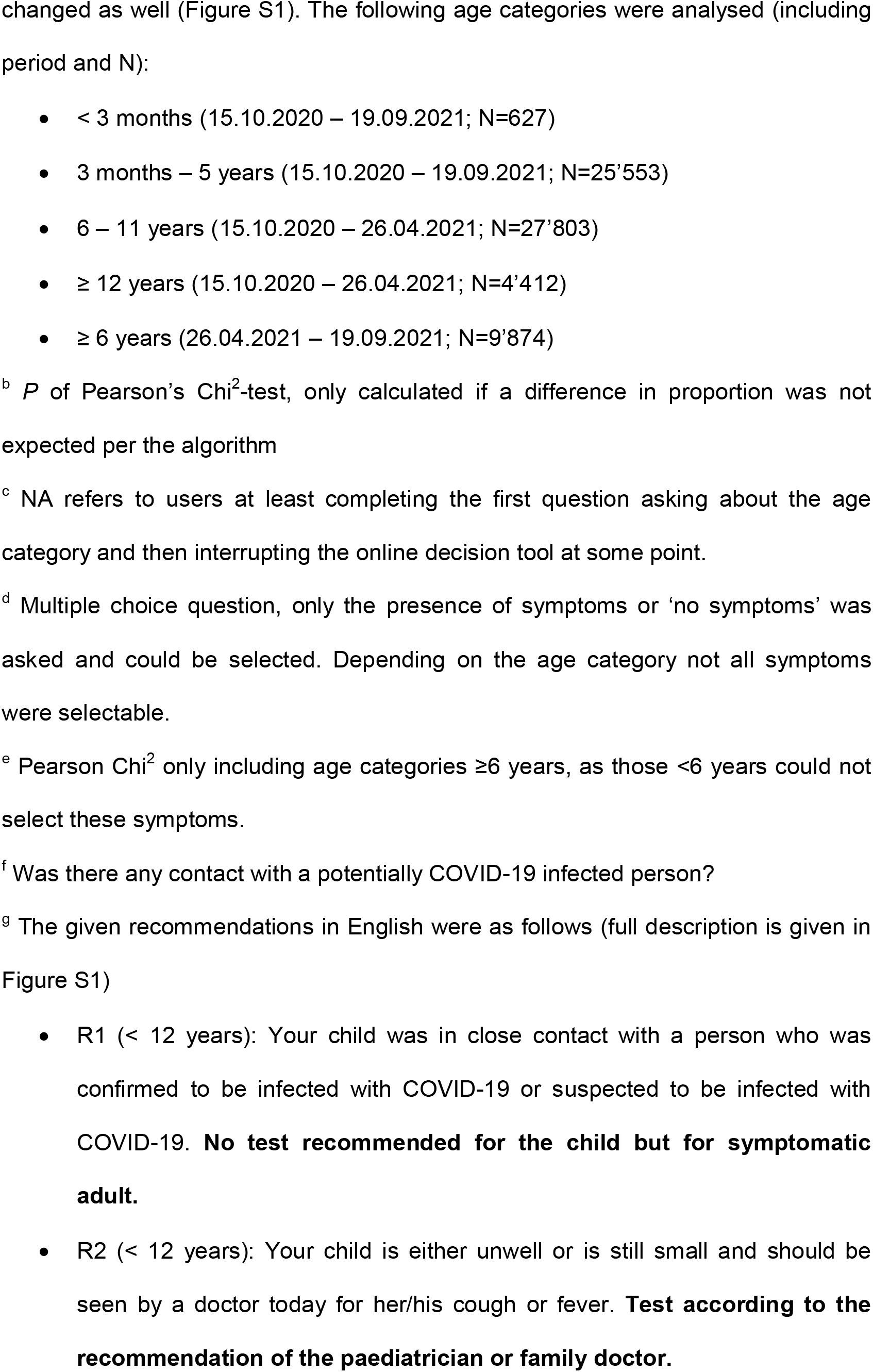
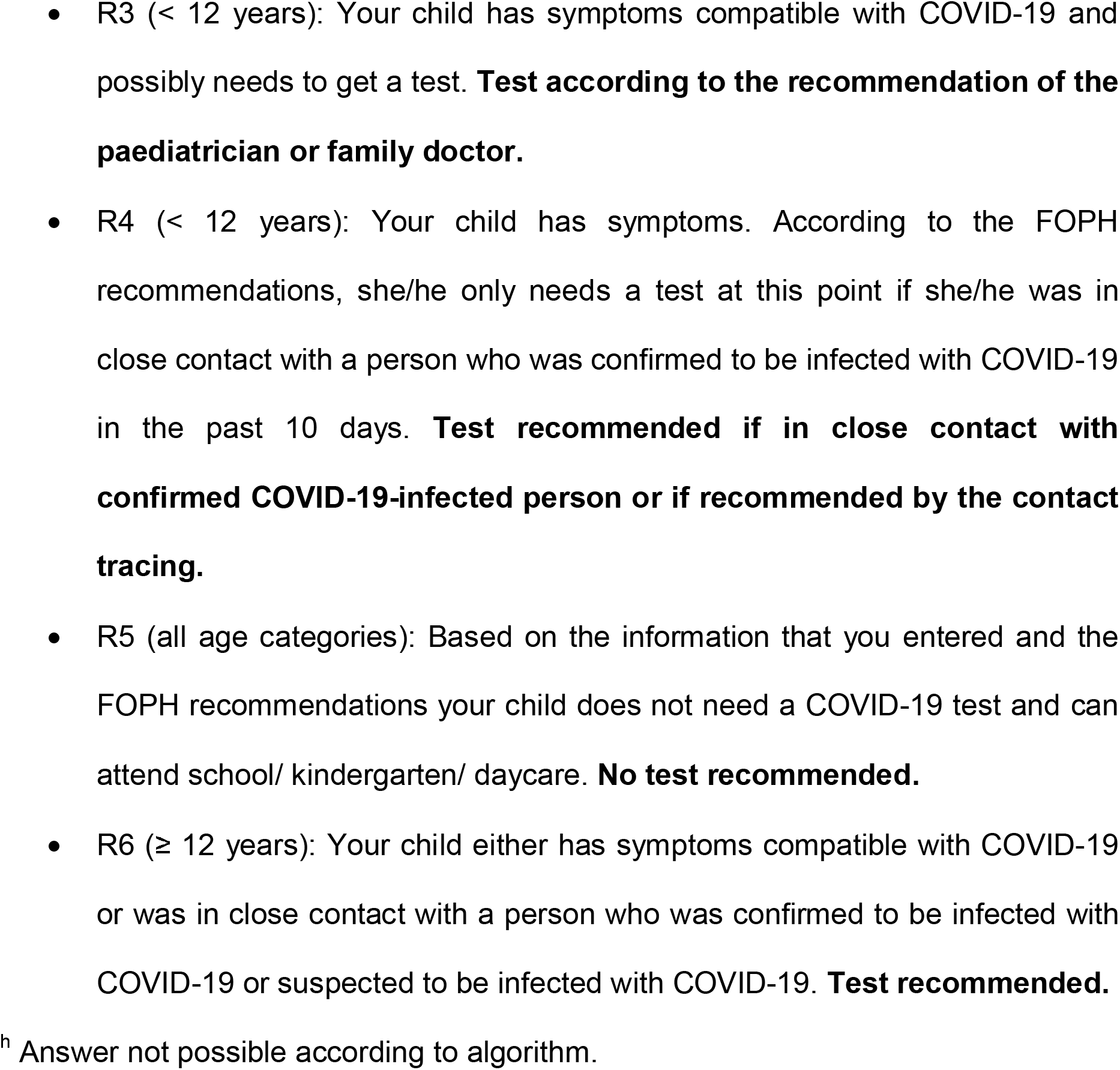
Findings from the online decision tool (N = 68’269)

15% (10’304/68’269) had close contact to a confirmed or suspected case of COVID-After the FOPH extended the adult testing recommendation (testing in case of any symptom) for children ≥6 years the proportion of users reporting a COVID-19 contact increased from 14% (7’133/50’896) to 18% (3’171/17’373), 𝒳^2^=181, *P*=<.001. Accordingly, fewer users reported any symptom after the change in testing strategy: 86% (43’958/50’896) versus 78% (13’573/17’373), 𝒳^2^=664, *P*=<.001. We observed a discrepancy in the proportion of reported contact with a confirmed or suspected case of COVID-19 (15% (10’304/68’269)) compared to the number of users that were contacted by the cantonal contact tracing (3% (2’382/68’269)). 60% (40’890/68’269) of surveys were filled in outside of regular office hours (Table 1). The Swiss testing recommendation was more restrictive for children <12 years (and <6 years as of April 2021) when compared to older children and adults. Accordingly, the tool recommended a test in 71% (18’648/26’180) of children <6 years, compared to 82% (34’356/42’089) of children ≥6 years, 𝒳^2^=1.0e+3, *P*=<.001.

### Voluntary fields, initial survey

There was no significant difference in the proportion of school-aged children (≥ 6 years) in the mandatory and voluntary survey: 62% (42’089/68’269) and 61% (21’319/35’068), respectively (𝒳^2^=7, *P*=.007). Age was the only field that was present in both the mandatory and the voluntary part of the online decision tool and could be used to assess representativeness of the voluntary fields. The majority of users were parents: 60% (12’278/20’330) were mothers and 32% (6’519/20’330) fathers. Only 1% (218/20’330) were healthcare professionals (Table 1). Most responders came from the Central Plateau (39% (7’992/20’496)), which inhabits 22% (1’904’451/8’717’105) of the Swiss population and also includes the canton of Bern [16].

### Follow-up survey

The follow-up survey was filled in by 1’399 users and contained questions around socio-economic status, the reasons for use, and the COVID-19-tests results (Table 2). We saw no significant difference in the proportion of school-aged children (≥6 years), sex, relation to the child, and geographic region between the voluntary fields from the online decision tool and the follow-up survey, suggesting representativeness (Table S4).

**Table 2.**
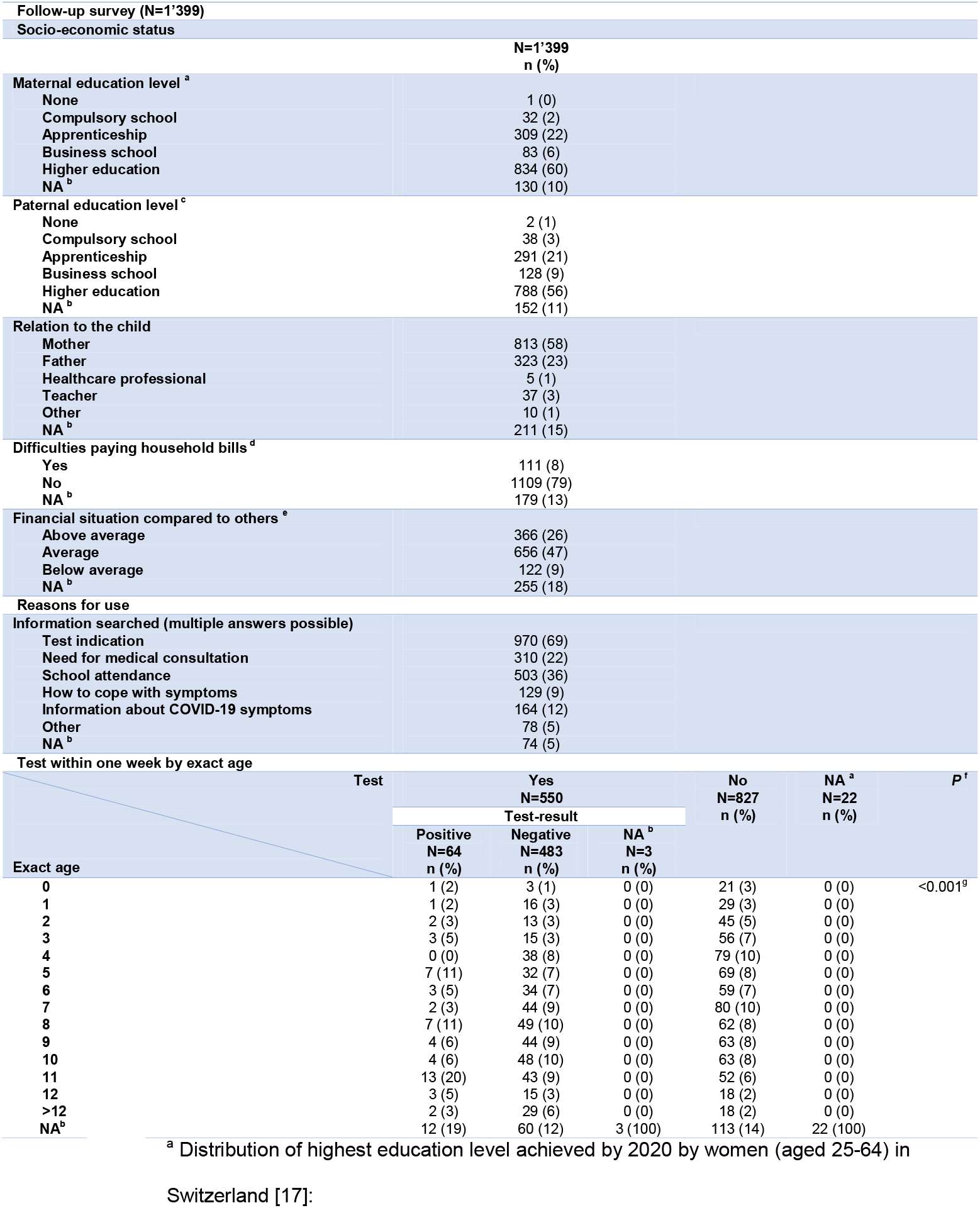
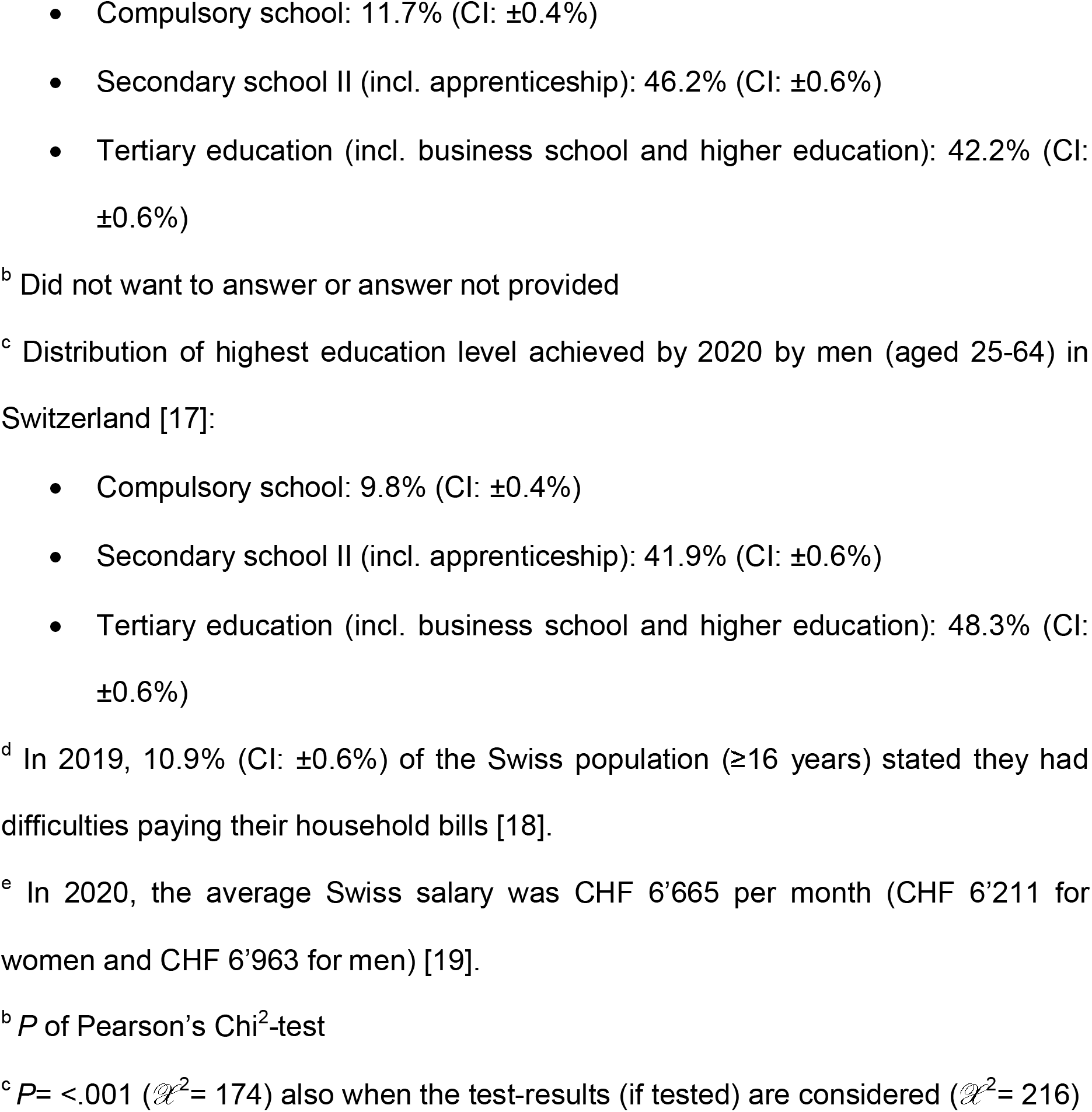
Data from the follow-up survey (N=1’399)

The majority of parents reported to have Higher Education (university, technical college): 60% (834/1’399) and 56% (788/1’399) of mothers and fathers, respectively. Most users reported a stable financial situation: 79% (1’109/1’399) responded that they had no difficulties paying their household bills. 47% (656/1’399) rated their financial situation as average and 26% (366/1’399) as being above the national average.

### Reasons for use and testing

Reasons for use are shown in Table 2. 88% (1’170/1’325) of respondents used coronabambini.ch for information that fell into the scope of the tool (need for testing and school attendance). 41% (546/1325) of users searched for additional information, including: need for a medical consultation (23% (310/1’325)), information on COVID-19 symptoms (12% (164/1’325)), and how to deal with symptoms (10% (129/1’325)).

30% (131/430) of children <6 years of age performed a COVID-19 test within one week of using coronabambini.ch (11% (14/131) tested positive), compared to 45% (344/759) and 11% (38/344) of children ≥ 6 years, respectively (Table 2).

### Adherence and reasons why

Information regarding adherence and reasons for adherence is displayed in Table 3. Overall adherence to the recommendation of the online tool was 84% (1’131/1’352), there was no difference in adherence when a test was possibly recommended (consult physician) or not recommended (observation or school attendance): 87% (434/499) and 83% (576/695) respectively, 𝒳^2^=4, *P*=.053. The highest adherence was observed when the tool recommended to stay at home and observe (89% (344/385)). The lowest adherence was observed when school attendance was recommended (75% (232/310)).

**Table 3.**
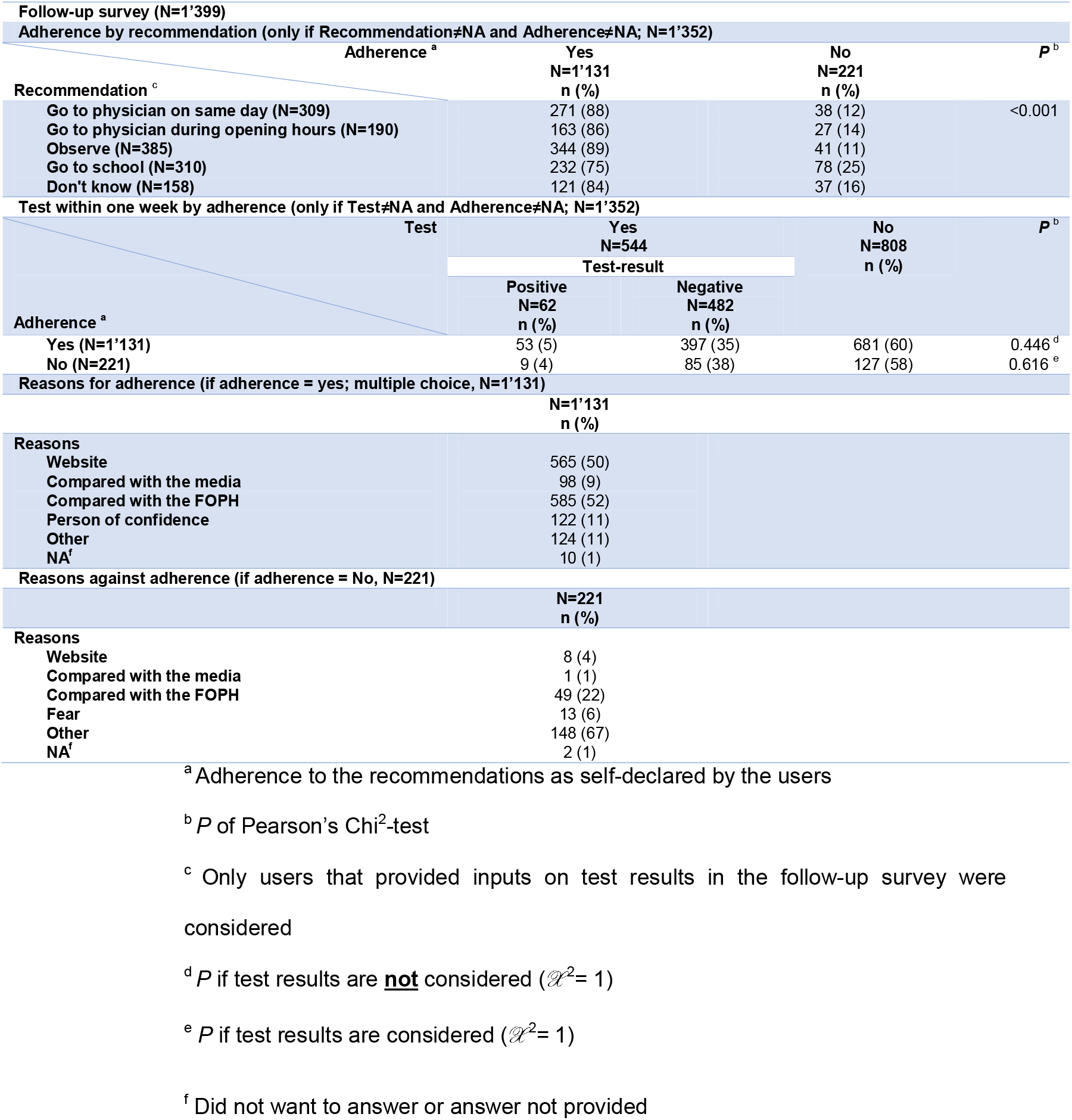
Adherence to recommendations of the online decision tool

Adherence to the recommendation was not different among children who finally tested positive (86% (53/62)) compared to those who tested negative (83% (397/482)), 𝒳^2^=0, *P*=.541 (Table 3).The main reason to follow the recommendation was that the results were compared with the FOPH guidelines (52% (585/1’131)). 22% (49/221) of non-adherers also compared the recommendation of the online tool to the FOPH guidelines.

## Discussion

### Main findings

We successfully implemented an online decision support tool, coronabambini.ch, to support caregivers, teachers and healthcare personnel to comply with the Swiss national pediatric testing strategy. Translation of a relatively complex testing guideline into a decision algorithm was possible and the tool has been kept up to date with current testing recommendations until it was discontinued. Given the more differentiated pediatric testing approach the design and implementation of the branches for smaller children was only just possible in a commercial web solution supporting simple branching and skip logic. Given the voluntary nature of the demographic and socioeconomic questions, assessment of the degree and representativeness of usage was not fully possible. In the Central Plateau area, where the tool was developed and promoted, usage was high: taking the information of users about the region, we estimate that about 26’600 (39% of 68’269) users came from the Central Plateau area. This number of users corresponds to approximately 7% (26’600/375’515) of the population under 20 years in this region [16]. The adherence with the tool’s recommendation was generally high. A large proportion of users reported that they compared the recommendation of the tool with other information sources, including the FOPH testing guideline. Almost one quarter of the non-adherers drew different conclusions from the FOPH guidelines when compared to the tool, pointing to the large variability in the interpretation of written guidelines. Algorithms, such as coronabambini.ch, reply on exact choices for each branching (Figure S1), contrary to written guidelines, which may be more vague and allow for a greater degree of user interpretation.

We saw reflection of some epidemiological characteristics of the COVID-19 pandemic in the information captured by coronabambini.ch. For example, gastrointestinal symptoms were reported more frequently in the lower age group, which has been well documented among children [20-23]. Over the summer months, when fewer other viral infections circulated in the community, fewer users consulted the tool because of signs of illness, and the proportion of users with a positive COVID-19 contact concurrently increased, compared to winter months. Further, we could show that the usage in general corresponded well to the course of the pandemic as seen in Figure 2. The additional peaks in usage, not correlating with the pandemic waves, could be explained by the school-holidays and the return to school attendance at each of these periods (December, February, April and August) as well as the update of the test algorithm (end of April). We were also able to detect differences in the acceptability of public health recommendations depending on the testing recommendation. Among the recommendations to not test immediately, watchful waiting at home showed greater acceptance compared to school or daycare attendance. These examples point to the potential of online decision tools for epidemiological surveillance and monitoring of policy implementation. In addition, online tools may hint to information gaps in the public health system.

However, our data from the tool and the follow-up survey was likely subject to significant selection bias. For example, it is likely that users with high digital literacy and higher education were more likely to use the tool and to respond to the follow-up survey. We were unable to fully measure the degree and type of bias in our data due to the inherent limitations of such a voluntary digital online tool. This represents an important and inherent methodological limitation, which has to be taken into account when considering such tools for public health surveillance.

In the case of coronabambini.ch, we observed a discrepancy in the proportion of reported contact with a confirmed or suspected case of COVID-19 compared to the number of users that were contacted by the cantonal contact tracing. A possible explanation could be that the positive-tested person has to report possible close contacts to the cantonal contact tracing; therefore, some might have not reported children to avoid quarantine requirements for them. Furthermore, coronabambini.ch may have filled in an information gap until families were contacted by telephone contact tracers. However, these explanations are only speculative since data on COVID-19 contact tracing is not available to us.

### Comparison with prior work

Mothers were the primary users of our online decision tool. Although it has been reported that men may have a higher level of adoption intention of e-health applications [24], it is most probable that women as caregivers were more likely to assume healthcare responsibilities [25]. This female gender predominance was also observed in the recent implementation of a COVID-19 online decision support tool for adults in Switzerland [14].

Approximately 41% (546/1325) of responders had additional expectations in opening the tool, including medical advice. This underlines the importance in communicating the scope of the tool, which also appeared as an important factor around usability of coronabambini.ch in a qualitative study including users of the tool [26]. The majority of users, regardless of adherence, compared the recommendations given by coronabambini.ch with other information sources such as the internet, the media or the FOPH-guidelines, which corresponds with the findings in another study [14]. Nonetheless, 84% (1’131/1’352) of users followed the instructions given by coronabambini.ch. That leads to the conclusion that a majority of them trusted our online decision tool. This corresponds with the findings of *Michel et al*. stating that the main reason for adherence was ‘trust in the online forward triage tool (OFTT) (40.3%)’ with an adherence of 84.7% [14]. Other studies reported similar adherence rates to OFTTs and teleconsultations in Switzerland [27-29]. The high rate of adherence to testing recommendations points to the potential of online decision tools in unburdening the health system, as described in other studies [4-6,14,30,31].

### Limitations

This analysis has important methodological limitations resulting from the design of coronabambini.ch as a public health, rather than a research tool. The low response rate of the follow-up survey is an important limitation of our analysis. Only a small proportion of users who answered all mandatory questions also filled in the follow-up survey. This may limit the robustness of some of the conclusions drawn from the results. The reason for the low response rate mainly lies in the voluntariness of providing an email address. Further, the request to participate was sent automatically and there was no reminder to participate. We can only roughly estimate the degree of selection bias in the follow-up survey as, for example, demographic data, was not obtained as part of the mandatory fields. It is likely that the level of adherence was overestimated due to the selection of favorable respondents in the follow-up survey. There was also an unusually high proportion of participants with Higher Education in the follow-up survey (55% of responders vs. 30% in the general population) [17]. This selection bias was unavoidable, as we wanted to create a free tool for the public and not primarily a research tool. We anticipate that any tool designed primarily intended for public health needs would have to deal with such biases when interpreting data for surveillance purposed and would require advanced imputation measures. This bias could have also influenced our findings regarding on comprehensibility. One study showed that adult users in an academic setting could correctly identify recommended care instructions from a self-triage website during a pandemic [5], but there is no data regarding the general population. Further, the scope of our decision support tool was very limited when compared to medical decision support tools for patients or health-care professionals [32,33]. Moreover, given the time lack between the initial and follow-up survey, there is a risk for recall bias in the follow-up survey. The Central Plateau region of Switzerland is overrepresented in our sample: 39% (7’992/20’496) of users were living in the Central Plateau, which also represents the most populated area in Switzerland. The online decision tool was developed and promoted in Bern, which explains the overrepresentation of the Central Plateau area.

### Online decision support tools: opportunities and challenges

The current COVID-19 pandemic is not the first pandemic in which e-health and telemedicine were used as an OFTT. Already during the H1N1-pandemic in 2009 these triage systems were introduced, e.g. the Strategy for Off-site Rapid Triage [34] or the MLN FluLine [30]. The COVID-19 pandemic further stimulated digitalization and telehealth [35]. Forward triage, including OFTT, are playing a central role in pandemics [31]. However, these systems were developed for adult users as there was concern that an interactive website might discourage some parents from contacting their child’s medical provider [34], although many parents seemed to use these services regardless [30]. When compared to more complex medical advice, clear-cut testing recommendations, as the guidelines implemented in www.coronobambini.ch, are easier to translate into trustable online decision support tools even for pediatric patients.

Other studies have already stated that self-triage telemedicine tools could prevent unnecessary emergency department (ED) visits as well as reducing potential infectious exposures and transmissions [4-6,14,30,31], not only during pandemics [28,29]. However, to make better use of the potential of such an online decision support tool, including public health surveillance, its complexity and performance should be improved [36-38]. Further, when using routine online tools for disease surveillance, policy makers should be well aware of the inherent selection biases of such tools. Efforts should be made to be able to measure the degree and type of bias, for example through estimating key demographic, socioeconomic and health literacy factors of users.

## Conclusion

Usage of coronabambini.ch was generally high. Acceptance of the tool was high, especially amongst people with higher education and high health literacy, and it was used more frequently in areas where it was developed and promoted.

Certain patterns in epidemiology and adherence to public health policy could be depicted but selection bias was difficult to measure showing the potential and challenges of digital decision support as public health tools in providing information on disease epidemiology.

## Supporting information

RECORD Checklist

## Data Availability

Statistical code and dataset are available upon reasonable request.

## Abbreviations

FOPH: Swiss Federal Office of Public Health
OFTT: online forward triage tool
ED: emergency department

## Author contributions

Conception/design: CA, IS, FR, TCS, KK, CS and NT

Development of coronabambini.ch/data acquisition: CA, FR, TCS, KK, CS, IS and NT

Data analysis/interpretation: CS, NT, TCS and KK

Manuscript draft: CS, NT, TCS and KK

All authors critically revised and approved the final version of the manuscript.

## Acknowledgements

Aristomenis Exadaktylos, Department of Emergency Medicine, Inselspital, Bern University Hospital, University of Bern, Switzerland: support for the development and evaluation of coronabambini.ch

Linda Adamikova, Bettina Ley and Brigitte Meier at the FOPH: feedback on the algorithm, critical revision of the manuscript.

## Reporting statement

RECORD/STROBE-checklist available separately

## Patient consent form

Not applicable

## Ethics approval

The local ethics committee of the Canton of Bern, Switzerland deemed our study exempt from full ethical approval (REQ-2020-01179). Though full informed consent was hence not required, we did add an information on the first page of coronabambini.ch that data entered in the tool may be used for research at the University of Bern (Figure S2).

## Data sharing statement

Statistical code and dataset are available upon reasonable request.

## Figure legend

Figure S1. Logic-tree for coronabambini.ch. S1A: Version 1 (15 October 2020); S1B: Version 2 as per updated FOPH testing guidelines (26 April 2021).

Figure S1B.

Figure S2. Screenshot of landing page and first question of coronabambini.ch (English version).

