## Supplementary figures and images for "coronabambini.ch: Development and usage of an online decision support tool for pediatric COVID-testing in Switzerland: a cross-sectional analysis"

### Supplementary Figures

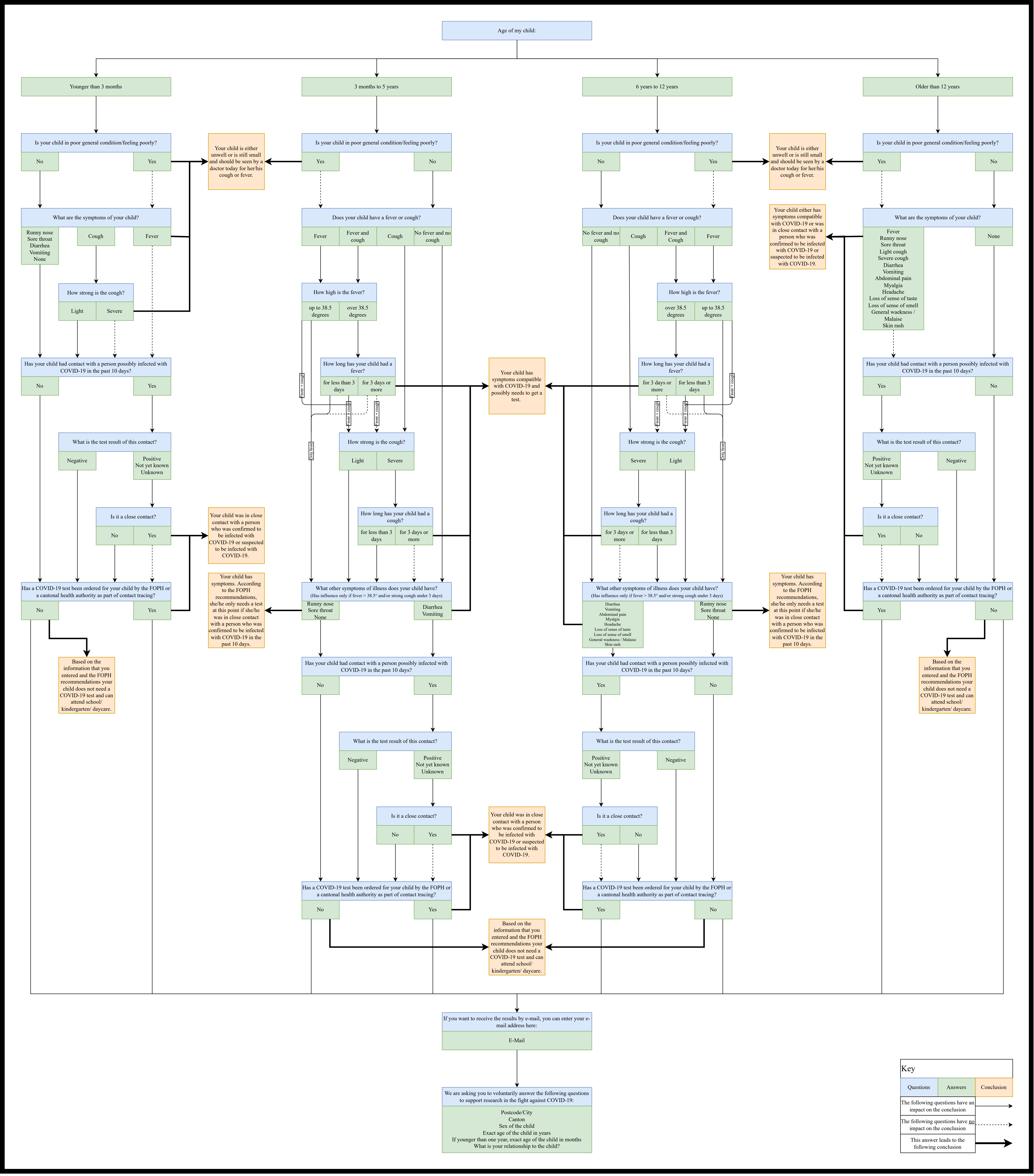

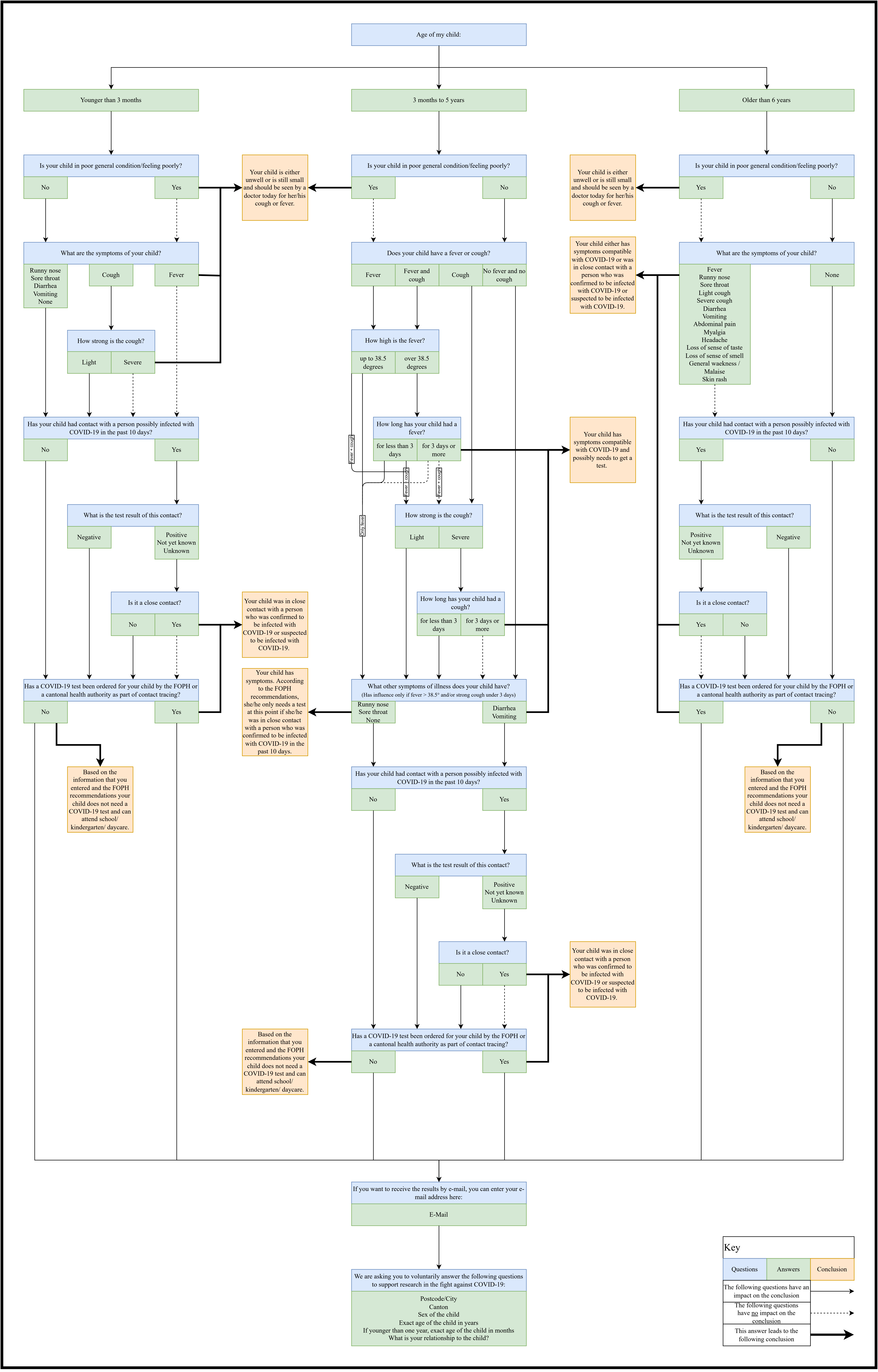

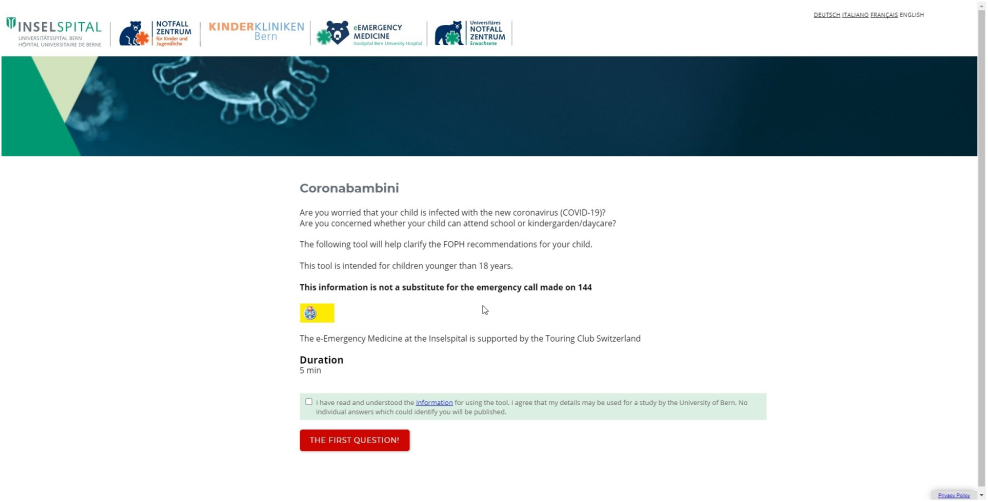

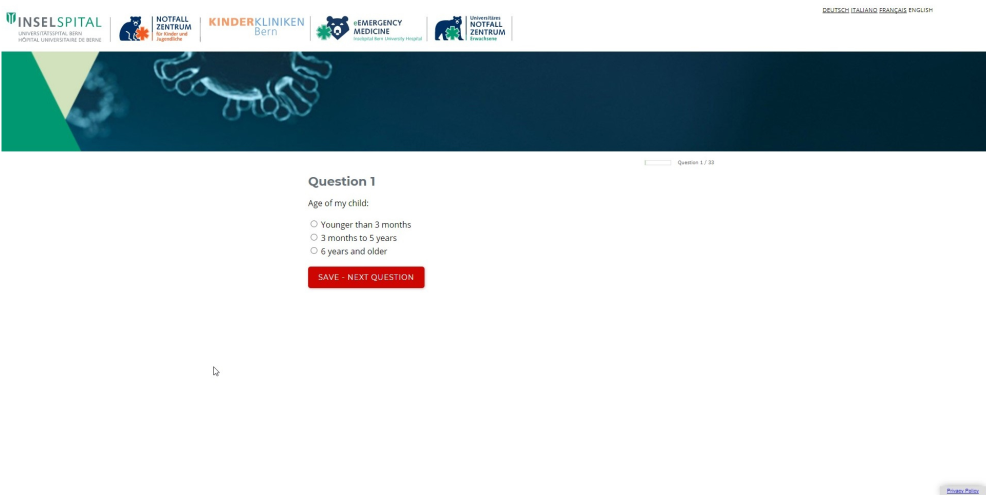
