## Supplementary Tables for "coronabambini.ch: Development and usage of an online decision support tool for pediatric COVID-testing in Switzerland: a cross-sectional analysis"

Table S1. Mandatory and voluntary questions (including answers) of www.coronabambini.ch.

| Number [a] | Question and answers |
|---|---|
| **1** | **Age of my child:** |
| | Younger than 3 months |
| | 3 months to 5 years |
| | 6 years to 12 years |
| | Older than 12 years |
| **2** | **Is your child in poor general condition/feeling poorly?:** |
| | Yes |
| | No |
| **3** | **What are the symptoms of your child?:** |
| | Fever |
| | Runny nose |
| | Sore throat |
| | Cough |
| | Diarrhoea |
| | Vomiting |
| | None |
| **3.1** | **How strong is the cough?:** |
| | Light |
| | Severe |
| **4** | **Does your child have a fever or cough?:** |
| | Fever |
| | Cough |
| | No fever and no cough |
| | Fever and cough |
| **4.1** | **How high is the fever?:** |
| | Up to 38.5 degrees |
| | Over 38.5 degrees |
| **4.1.1** | **How long has your child had a fever?:** |
| | For less than 3 days |
| | For 3 days or more |
| **4.2** | **How strong is the cough?:** |
| | Light |
| | Severe |
| **4.2.1** | **How long has your child had a cough?:** |
| | For less than 3 days |
| | For 3 days or more |
| **5** | **What other symptoms of illness does your child have?** |
| | Runny nose |
| | Sore throat |
| | Diarrhoea |
| | Vomiting |
| | None |
| **6** | **What other symptoms of illness does your child have?** |
| | Runny nose |
| | Sore throat |
| | Diarrhoea |
| | Vomiting |
| | Abdominal pain |
| | Myalgia |
| | Headache |
| | Loss of sense of taste |
| | Loss of sense of smell |
| | None |

### Table S1. continued

| | |
|---|---|
| **7** | **What are the symptoms of your child?**<br>Fever<br>Runny nose<br>Sore throat<br>Light cough<br>Severe cough<br>Diarrhoea<br>Vomiting<br>Abdominal pain<br>Myalgia<br>Headache<br>Loss of sense of taste<br>Loss of sense of smell<br>General weakness / Malaise<br>Skin rash<br>None |
| **8** | **Has your child had contact with a person possibly infected with COVID-19 in the past 10 days?**<br>Yes<br>No |
| **8.1** | **What is the test result of this contact?**<br>Positive<br>Negative<br>Not yet known<br>Unknown |
| **8.1.1** | **Is it a close contact?**<br>Yes<br>No |
| **9** | **Has a COVID-19 test been ordered for your child by the FOPH or a cantonal health authority as part of contact tracing?**<br>Yes<br>No |
| **10** | **If you would like to receive the results by mail and agree to be contacted once for a follow-up survey, you can enter your e-mail address here:**<br>Free text answer |
| **11** | **We are asking you to voluntarily answer the following questions to support research in the fight against COVID-19. If you prefer not to provide any information, you can simply skip the question and continue clicking without affecting the outcome:** |
| **12.1** | **Postcode:**<br>Free text answer |
| **12.2** | **City:**<br>Free text answer |
| **12.3** | **Canton:**<br>Zurich<br>Berne<br>Lucerne<br>Uri<br>Schwyz<br>Obwald<br>Nidwald<br>Glarus<br>Zoug<br>Friburg<br>Soleure<br>Basle-City<br>Basle-Country<br>Schaffhouse<br>Appenzell Outer-Rhodes<br>Appenzell Inner-Rhodes<br>St. Gall<br>Grisons<br>Argovia<br>Thurgovia<br>Ticino<br>Vaud<br>Wallis<br>Neuchâtel<br>Geneva<br>Jura |

Table S1. continued

| | |
|---|---|
| **12.4** | **Sex of the child:**<br>Male<br>Female |
| **12.5** | **Exact age of the child in years:**<br>0<br>1<br>2<br>3<br>4<br>5<br>6<br>7<br>8<br>9<br>10<br>11<br>12<br>>12 |
| **12.6** | **If younger than one year, exact age of the child in months**<br>1<br>2<br>3<br>4<br>5<br>6<br>7<br>8<br>9<br>10<br>11 |
| **12.7** | **What is your relationship to the child?:**<br>Mother<br>Father<br>Teacher<br>Healthcare professional<br>Other |

[a] The following questions were asked for each age category:

- < 3 months (15.10.2020 – 16.02.2022): 1 – 3, 8 – 12
- 3 months – 5 years (15.10.2020 – 16.02.2022): 1, 2, 4, 5, 8 – 12
- 6 – 11 years (15.10.2020 – 26.04.2021): 1, 2, 4, 6, 8 – 12
- ≥ 12 years (15.10.2020 – 26.04.2021): 1, 2, 7 – 12
- ≥ 6 years (26.04.2021 – 16.02.2022): 1, 2, 7 – 12

Table S2. Voluntary questions (including answers) of the follow-up survey.

| Number | Question and answers |
|---|---|
| **1** | **What did you do after using www.coronabambini.ch?:**<br>I took my child to the emergency department.<br>I called my pediatrician/family doctor the same day.<br>My pediatrician/family doctor's office was closed and I called him/her the next days during normal business hours next day.<br>I visited my pediatrician's/family doctor's office without calling ahead of time.<br>I kept my child at home but did not call my pediatrician/family doctor.<br>I sent my child to school/daycare.<br>Something else |
| **1.1** | **What exactly did you end up doing?:**<br>Free text answer |
| **2** | **Did your child get tested for COVID-19 within one week of filling in www.coronabambini.ch?:**<br>Yes<br>No |
| **2.1** | **What was the result of your child's COVID test?:**<br>Positive<br>Negative |
| **3** | **Did anybody from your household or close contacts get tested?:**<br>Yes, family<br>Yes, close contacts in school/Kindergarten/Daycare<br>No |
| **3.1** | **What was the result?:**<br>Positive<br>Negative |
| **4** | **Did your child require medical care within one week of filling in www.coronabambini.ch?:**<br>Yes, at a pediatrician/family doctor's office.<br>Yes, at the emergency department.<br>Yes, my child was admitted to the hospital.<br>No. |
| **5** | **What did www.coronabambini recommend you?:**<br>To contact our pediatrician/family doctor the same day.<br>To contact our pediatrician/family doctor within normal business hours.<br>To watch my child's state at home and to contact our pediatrician/family doctor in case of worsening or persistence of symptoms.<br>To send my child to school/kindergarten.<br>I don't remember |
| **6** | **Did you stick to the recommendations of www.coronabambini.ch?:**<br>Yes<br>No |
| **6.1** | **What made you follow the recommendation of www.coronabambini.ch?:**<br>I trust the website as a reliable information source.<br>I compared the recommendations with recommendations from the media and took a decision.<br>I compared the recommendations with those from the FOPH and took a decision.<br>I sought advice from a person I trusted.<br>Other |
| **6.1.1** | **What other reasons made you follow the recommendation of www.coronabambini.ch?:**<br>Free text answer |
| **6.2** | **What made you NOT follow the recommendation of www.coronabambini?:**<br>I did not trust the website as a reliable source of information.<br>The recommendations from the website differed from the media recommendations.<br>I compared the recommendations with those from the FOPH and took a decision.<br>I feared for my child and preferred to consult the paediatrician<br>Other |
| **6.2.1** | **What other reasons made you NOT follow the recommendation of www.coronabambini.ch?:**<br>Free text answer |
| **7** | **What information were you looking for? I wanted...:**<br>... to know if my child needs a test.<br>... to know if I need to consult my child's pediatrician/family doctor.<br>... to know if my child can go to school.<br>... more information on how to cope with COVID-19 symptoms.<br>... more information on COVID-19 symptoms.<br>... other |
| **7.1** | **What other information were you looking for?:**<br>Free text answer |
| **8** | **Did you find the information that you needed?:**<br>Yes.<br>No, because the information was not comprehensive.<br>No, because the information was not complete. |

Table S2. continued

| | |
|---|---|
| **9** | **How did you learn about www.coronabambini.ch?:**<br>Through my pediatrician/family doctor<br>Through the internet/media<br>Through my school<br>Through friends/family<br>Through the FOPH website<br>I was redirected through the FOPH Coronachecker<br>Other |
| **9.1** | **From what other source did you hear about www.coronabambini.ch?:**<br>Free text answer |
| **10** | **Did www.coronabambini.ch adequately address your concerns regarding COVID-19?:**<br>Yes, the information from the website reassured me.<br>No, the information from the website did not reassure me.<br>No, the information from the website increased my fears and anxieties.<br>I was not worried in the first place. |
| **11** | **How did you cope with your fears? What helped you cope?:**<br>Free text answer |
| **12** | **What is the highest education degree achieved by the child's mother?:**<br>Did not go to school<br>Mandatory education (or a few years at school)<br>Apprenticeship<br>Higher technical or commercial college<br>University<br>I do not know or do not want to answer |
| **13** | **What is the highest education degree achieved by the child's father?:**<br>Did not go to school<br>Mandatory education (or a few years at school)<br>Apprenticeship<br>Higher technical or commercial college<br>University<br>I do not know or do not want to answer |
| **14** | **Did you have difficulties paying your household bills during the last 12 months?:**<br>Yes<br>No<br>I do not know or do not want to answer |
| **15** | **Compared to other families in Switzerland, the financial situation of your family is...:**<br>...above average<br>...average<br>...below average<br>...I do not know or do not want to answer |
| **16** | **Is there anything general you want to tell us about www.coronabambini.ch?:**<br>Free text answer |
| **17** | **In a second stage, we will ask individual participants of this survey at a later date to take part in individual interviews for an expense compensation in order to gain further knowledge about the use of online triage tools. If you agree, we ask you to leave your contact number. The number will only be used in the context of this survey and will never be passed on.:**<br>Yes, I consent to be contacted: free text answer<br>No, please, no more interviews |
| **18** | **Due to the guarantee of anonymity, your survey information from www.coronabambini.ch has not been linked to your email. We ask you to fill in this information again. Thank you very much:** |
| **18.1** | **Postcode:**<br>Free text |
| **18.2** | **City:**<br>Free text |

Table S2. continued

| | |
|---|---|
| **18.3** | **Canton:**<br>Zurich<br>Berne<br>Lucerne<br>Uri<br>Schwyz<br>Obwald<br>Nidwald<br>Glarus<br>Zoug<br>Friburg<br>Soleure<br>Basle-City<br>Basle-Country<br>Schaffhouse<br>Appenzell Outer-Rhodes<br>Appenzell Inner-Rhodes<br>St. Gall<br>Grisons<br>Argovia<br>Thurgovia<br>Ticino<br>Vaud<br>Wallis<br>Neuchâtel<br>Geneva<br>Jura |
| **18.4** | **Sex of the child:**<br>Male<br>Female |
| **18.5** | **Exact age of the child in years:**<br>0<br>1<br>2<br>3<br>4<br>5<br>6<br>7<br>8<br>9<br>10<br>11<br>12<br>>12 |
| **18.6** | **If younger than one year, exact age of the child in months**<br>1<br>2<br>3<br>4<br>5<br>6<br>7<br>8<br>9<br>10<br>11 |
| **18.7** | **What is your relationship to the child?:**<br>Mother<br>Father<br>Teacher<br>Healthcare professional<br>Other |

Table S3. Comparison of symptoms given by parents (mother or father) or other users (healthcare professional, teacher or other) (N=20’330)

| **Online decision tool, mandatory fields (N=20’330)** | | | | | | | | | | | | | |
|---|---|---|---|---|---|---|---|---|---|---|---|---|---|
| **Symptoms by relation to the child and age category** | | | | | | | | | | | | | |
| **Relation to the child** | **Parents (N=18’797)** | | | | | | **Others (N=1’533)** | | | | | | |
| **Age category / Symptoms** | **< 3 months N=91 n (%)** | **3 months – 5 years N=6’932 n (%)** | **6 – 11 years N=8’144 n (%)** | **≥ 12 years N=911 n (%)** | **≥ 6 years N=2’719 n (%)** | **Total N=18’797 n (%)** | **< 3 months N=14 n (%)** | **3 months – 5 years N=509 n (%)** | **6 – 11 years N=701 n (%)** | **≥ 12 years N=113 n (%)** | **≥ 6 years N=196 n (%)** | **Total N=1’533 n (%)** | ***P*** **[a]** |
| **Ill appearing** | | | | | | | | | | | | | 0.921 |
| **Yes** | 19 (21) | 1044 (15) | 1025 (13) | 95 (10) | 286 (11) | 2’469 (13) | 4 (29) | 73 (14) | 93 (13) | 14 (12) | 16 (8) | 200 (13) | |
| **no** | 72 (79) | 5888 (85) | 7119 (87) | 816 (90) | 2433 (89) | 16’328 (87) | 10 (71) | 436 (86) | 608 (87) | 99 (88) | 180 (92) | 1333 (87) | |
| **Any symptoms [b]** | | | | | | | | | | | | | <0.001 |
| **Yes** | 74 (81) | 6262 (90) | 7548 (93) | 880 (97) | 2404 (88) | 17168 (91) | 12 (86) | 440 (86) | 621 (89) | 110 (97) | 165 (84) | 1348 (88) | |
| **No** | 17 (19) | 670 (10) | 596 (7) | 31 (3) | 315 (12) | 1629 (9) | 2 (14) | 69 (14) | 80 (11) | 3 (3) | 31 (16) | 185 (12) | |
| **Fever** | 30 (33) | 2855 (41) | 2370 (29) | 136 (15) | 666 (24) | 6057 (32) | 5 (36) | 199 (39) | 199 (28) | 22 (19) | 42 (21) | 467 (30) | 0.156 |
| **Cough** | 51 (56) | 3839 (55) | 3252 (40) | 286 (31) | 1175 (43) | 8603 (46) | 5 (36) | 283 (56) | 289 (41) | 32 (28) | 78 (40) | 687 (45) | 0.471 |
| **Runny nose** | 56(61) | 4046 (58) | 3911 (48) | 396 (33) | 1397 (51) | 9806 (52) | 9 (6) | 268 (53) | 283 (40) | 37 (33) | 81 (41) | 678 (44) | <0.001 |
| **Sore throat** | 10 (11) | 1262 (18) | 3219 (40) | 409 (45) | 819 (30) | 5719 (30) | 2 (14) | 96 (19) | 263 (38) | 51 (45) | 56 (29) | 468 (31) | 0.933 |
| **Diarrhoea** | 8 (9) | 498 (7) | 483 (6) | 54 (6) | 103 (4) | 1146 (6) | 3 (21) | 31 (6) | 41 (6) | 15 (13) | 13 (7) | 103 (7) | 0.329 |
| **Vomiting** | 5 (5) | 611 (9) | 547 (7) | 48 (5) | 158 (6) | 1369 (7) | 2 (14) | 49 (10) | 56 (8) | 8 (7) | 12 (6) | 127 (8) | 0.149 |
| **Abdominal pain** | - | - | 1470 (18) | 141 (15) | 308 (11) | 1921 (16) [c] | - | - | 94 (13) | 24 (21) | 15 (8) | 133 (16) [d] | 0.838 [e] |
| **Myalgia** | - | - | 595 (7) | 82 (9) | 111 (4) | 790 (7) [c] | - | - | 37 (5) | 7 (6) | 7 (4) | 51 (6) [d] | 0.534 [e] |
| **Headache** | - | - | 2514 (31) | 291(22) | 587 (22) | 3392 (29) [c] | - | - | 165 (24) | 33 (29) | 32 (16) | 230 (28) [d] | 0.513 [e] |
| **Loss of sense of smell** | - | - | 75 (1) | 22 (2) | 18 (1) | 115 (1) [c] | - | - | 9 (1) | 5 (4) | 0 (0) | 14 (2) [d] | 0.049 [e] |
| **Loss of sense of taste** | - | - | 107 (1) | 21 (2) | 29 (1) | 157 (1) [c] | - | - | 9 (1) | 3 (3) | 2 (1) | 14 (2) [d] | 0.393 [e] |
| **General weakness / Malaise** | - | - | 199 (2) | 139 (15) | 397 (15) | 737 (6) [c] | - | - | 22 (3) | 10 (9) | 19 (10) | 51 (6) [d] | 0.902 [e] |
| **Skin rash** | - | - | 33 (<1) | 8 (1) | 26 (1) | 67 (1) [c] | - | - | 5 (1) | 2 (2) | 4 (2) | 11 (1) [d] | 0.007 [e] |

[a] *P* of Pearson’s Chi$^2$-test, the “Total” data of each category (parents vs. others) was used to calculate the *P*-value.

[b] Multiple choice question, only the presence of symptoms or ‘no symptoms’ was asked and could be selected. Depending on the age category not all symptoms were selectable.

[c] Percentage calculated only including age categories ≥6 years, as those <6 years could not select these symptoms (N=11’774).

[d] Percentage calculated only including age categories ≥6 years, as those <6 years could not select these symptoms (N=829).

[e] Pearson Chi$^2$ only including age categories ≥6 years, as those <6 years could not select these symptoms (N=12’603)

Table S4. Comparison of demographic data entered by users of the online decision tool (voluntary fields) and the follow-up survey.

| **Source** | | Online decision tool n (%) | Follow-up-survey n (%) |
|---|---|---|---|
| **Age group** | | N=20’559 | N=1’189 |
| | <6 years | 7’464 (36) | 430 (36) |
| | ≥6 years | 13’095 (64) | 759 (64) |
| **Sex of child** | | N=19’658 | N=1’159 |
| | Female | 11’046 (56) | 658 (57) |
| | Male | 8’612 (44) | 501 (43) |
| **Responders** | | N=20’330 | N=1’188 |
| | Parents | 18’797 (92) | 1’136 (96) |
| | Others | 1’533 (8) | 52 (4) |
| **Regions** | | N=20’496 | N=1’200 |
| | Lake Geneva region | 1196 (6) | 98 (8) |
| | Central Plateau | 7992 (39) | 563 (47) |
| | Northwestern Switzerland | 2908 (14) | 154 (13) |
| | Zürich | 3317 (16) | 151 (13) |
| | Eastern Switzerland | 1629 (8) | 81 (7) |
| | Central Switzerland | 3206 (16) | 135 (11) |
| | Ticino | 248 (1) | 18 (2) |
